# Prediction of Metastatic Site Based On Somatic Gene Mutations in Primary Tumors in Prostate Cancer

**DOI:** 10.1101/2023.12.01.23299298

**Authors:** Paul Gomez

**Affiliations:** Nanobiotek, LLC

**Keywords:** Cancer, prostate cancer, metastasis, carcinoma, adenocarcinoma, data science

## Abstract

**Objective:** The goal of this research is to predict the most likely metastatic site(s) of a primary prostate cancer tumor that has been resected via radical prostatectomy; its genome has been sequenced to obtain a list of gene mutations; and after initial inspection of pelvic lymph nodes, there is no clinical evidence of metastasis. However, micrometastases might already be growing in distant organs and cannot be detected at the time of surgery.

**Background:** The most common metastatic targets in prostate cancer (PCa) are the pelvic lymph nodes (PLN) and bones. The PLNs are routinely dissected by a procedure called pelvic lymph node dissection (PLND) simultaneously with the surgical removal of the prostate to detect the presence of metastatic growths. Additionally, the prostate-specific antigen (PSA) level is used to assess the existence of a metastatic stage. However, micrometastases in other organs and tissues might be overlooked.

**Methods:** We downloaded publicly available prostate cancer tumor data from the website www.CbioPortal.org.

After choosing the 25 most commonly mutated genes by metastatic site (MS) and finding genes that are uniquely mutated on specific metastatic sites, we found that the mutational signature of a prostate cancer tumor is associated with its MS, and thus, we developed a method to numerically predict this association.

**Results:** After executing a computational algorithm on the data set of metastatic prostate tumors, it was found that we can predict metastatic sites with the following accuracies: bone (90.9%), retroperitoneum (87.5%), liver (83.0%), kidney (80.0%), pancreas (80.0%), adrenal glands (75.0%), lung (71.1%), and brain (72.5%).

**Conclusions:** We successfully developed a method and an algorithm that predict the most likely metastatic site of a primary prostate cancer tumor based on its genetic mutations. The accuracy of the predictions for eight metastatic sites ranges from 71.1% to 90.9%, with an average of 80.5%.

## INTRODUCTION

Prostate cancer (PCa) metastasizes to a variety of tissues and organs, with the pelvic lymph nodes (PLN) and bones being the most common sites [1][2][3]. During radical prostatectomy (RP) the PLN are dissected to check for metastatic lesions. This procedure is called pelvic lymph node dissection (PLND), of which there are several types. The American Urologic Association and the European Association of Urology recommend the extended PLND (EPLND) which includes the obturator, external, and internal iliac nodes [4]. PLND is almost always performed in cases when the probability of Lymph Node Involvement (LNI) is greater than 5% [5]. The risk of LNI is assessed by measuring the serum levels of prostate-specific antigen (PSA). The PSA level that is used as a threshold for bone metastasis is 100ng/mL [6][7]. Since the LN are always tested for metastasis, our algorithm does not use the LN as a predicted MS.

Besides the LN, PCa metastasizes preferentially to the bones [8][9][10]. Several imaging techniques exist to visualize bone metastases, namely, Whole-Body magnetic resonance imaging (WB-MRI) [11], positron emission tomography/computer tomography (PET/CT) [12], and others. With WB-MRI, a standard protocol has been established that targets the 14 most frequent metastatic sites (MS) identified on the Metastasis Reporting and Data System for Prostate Cancer (MET-RADS-P) template [13] shown on Figure 1.

**Figure 1.**
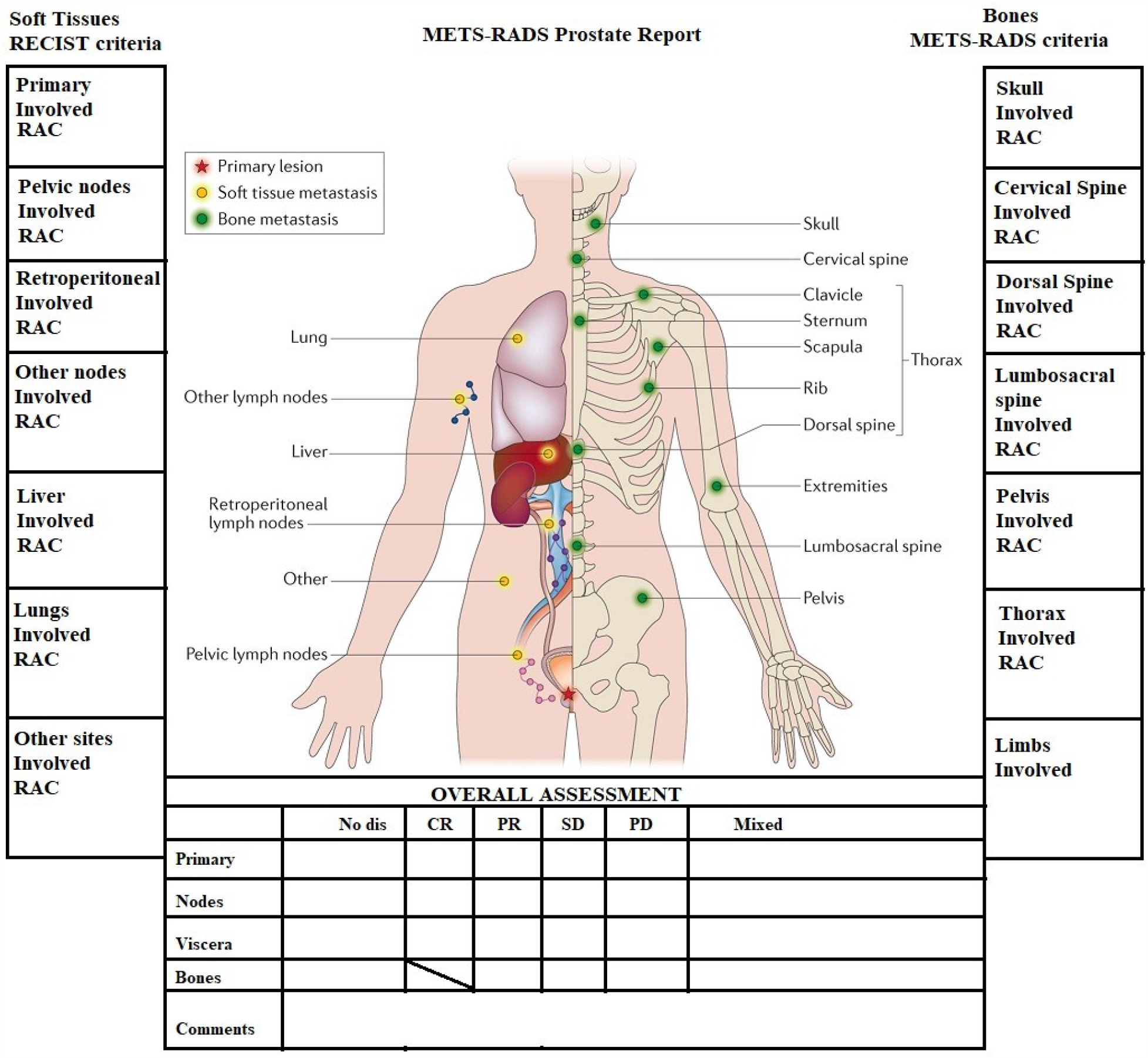
MET-RADS-P template showing the 14 most frequent metastatic sites

It should be noted that the brain and other less frequent metastatic sites, such as the spleen, pancreas, and the adrenal glands, are not included on the template. They all fall under the category “Other sites”. Those less frequent MS cases combined are less than 5% of all cases in our study but correspond to 47 patients, who could probably be saved by tailoring the WB-MRI to image specific organs should our predicting tool indicates a high probability of metastasis to those sites.

However, in many instances, no metastatic lesions are found after performing a WB-MRI. The main reason is that, although there might be micrometastases growing, their sizes are so small that they cannot be detected at the time of imaging.

We developed a computational tool that predicts the existence of metastasis in specific organs based on the mutations found in the primary tumor. Not finding clinical evidence of metastasis after RP should not mean that metastasis can be completely ruled out. Overlooking those MS could lead to unfavorable outcomes in the near future.

## MATERIALS AND METHODS

### Data Analysis

We downloaded prostate adenocarcinoma (PRAD) tumor data from the website www.CbioPortal.org [14], which holds cancer samples for all types of cancer. The data have been shared by several institutions and are publicly available. Using database methods, we cleaned up the data and selected all metastatic prostate cancer samples for detailed examination.

After choosing the 25 most commonly mutated genes by metastatic site and finding genes that are uniquely mutated on specific metastatic sites, we found that the mutational signature of a prostate cancer tumor is associated with its metastatic site, and thus, we developed a method to numerically predict this association.

The computer technology we used was Microsoft SQL Server 2022 to host the data and implement the prediction algorithm, and Python 3.10 (64-bit) to import the data into the SQL database.

We started with 1,758 metastatic prostate tumors, of which 636 metastasized to the lymph nodes. We excluded these tumors, as it was explained before. This study also excluded these other organs: the skin, muscles, penis, thorax, neck, urethra, and rectum, due to the small number of cases (<=3).

Additionally, the spleen (n=10) and the bladder (n=7) had to be excluded due to poor prediction accuracy (20% and 42%, respectively).

The final list of 8 metastatic sites we worked with for predicting the most likely MS is shown on Table 1, and graphically on the pie chart of Figure 2.

**Table 1.** Metastatic Sites in Prostate Cancer excluding LN.

| Metastatic site | Cases (n) | Percentage |
| --- | --- | --- |
| Bone | 506 | 59.7% |
| Liver | 218 | 25.7% |
| Lung | 76 | 9.0% |
| Brain | 16 | 1.9% |
| Kidney | 10 | 1.2% |
| Adrenal Gland | 8 | 0.9% |
| Retroperitoneum | 8 | 0.9% |
| Pancreas | 5 | 0.6% |

**Figure 2.**
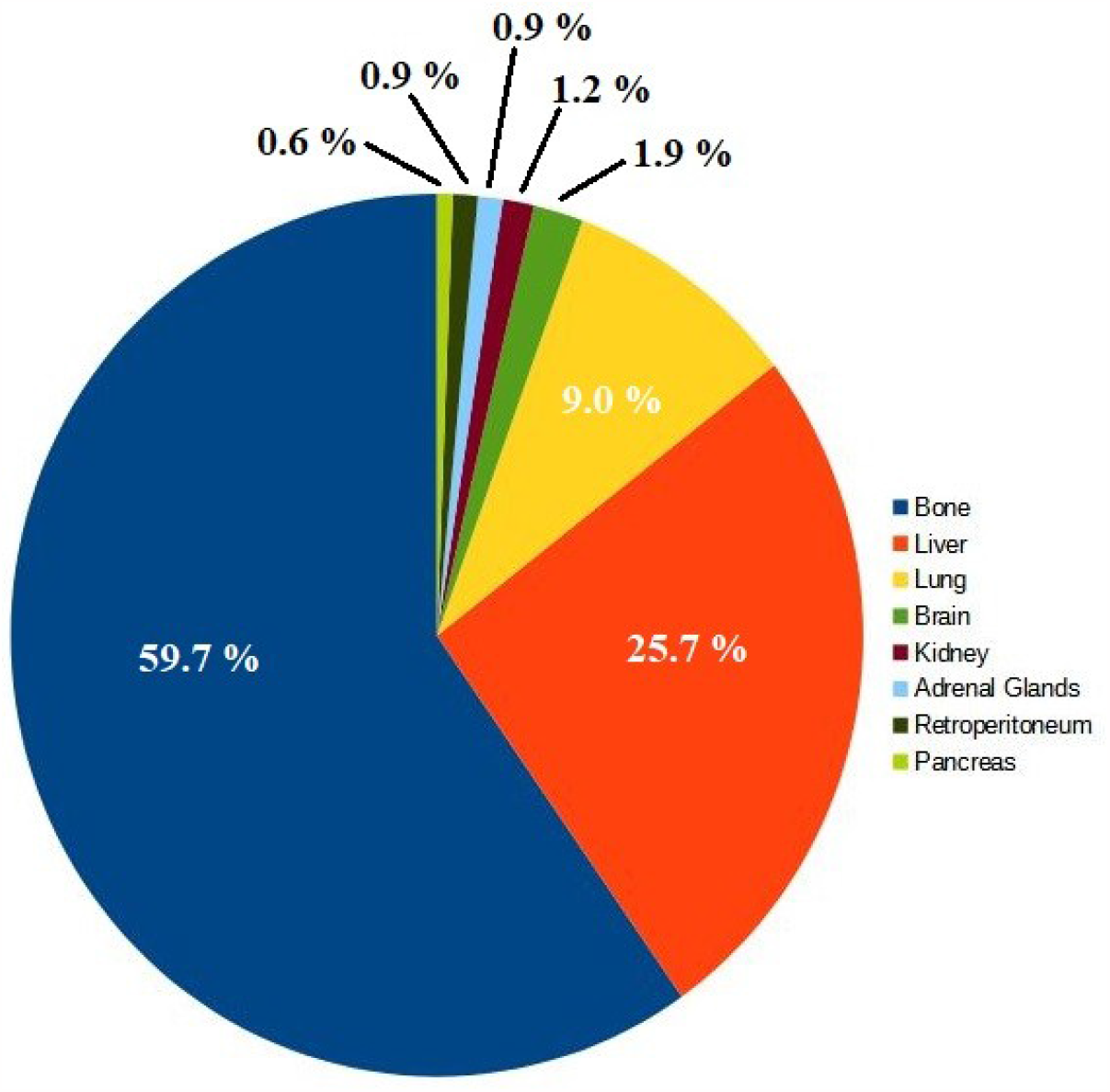
Metastatic Sites Cases in Prostate Cancer

A total of 847 metastatic tumors were used for further analysis. It can be seen from Table 1 and Figure 2 that the number of cases in which PCa metastasizes to the brain, kidney, adrenal glands, retroperitoneum, and pancreas is very small. However, when we analyzed the most commonly mutated genes by MS, it was discovered that they vary substantially depending on the metastatic site. This fact allowed us to design an algorithm based on competition.

The first step in the analysis of somatic mutations in PRAD neoplasms was to obtain the frequency of mutations in the top 25 most mutated genes. The rate of mutations was split into primary and metastatic genes, and then a list from an external source [15] was used for comparison. The results of this analysis are shown in Table 2.

**Table 2.**
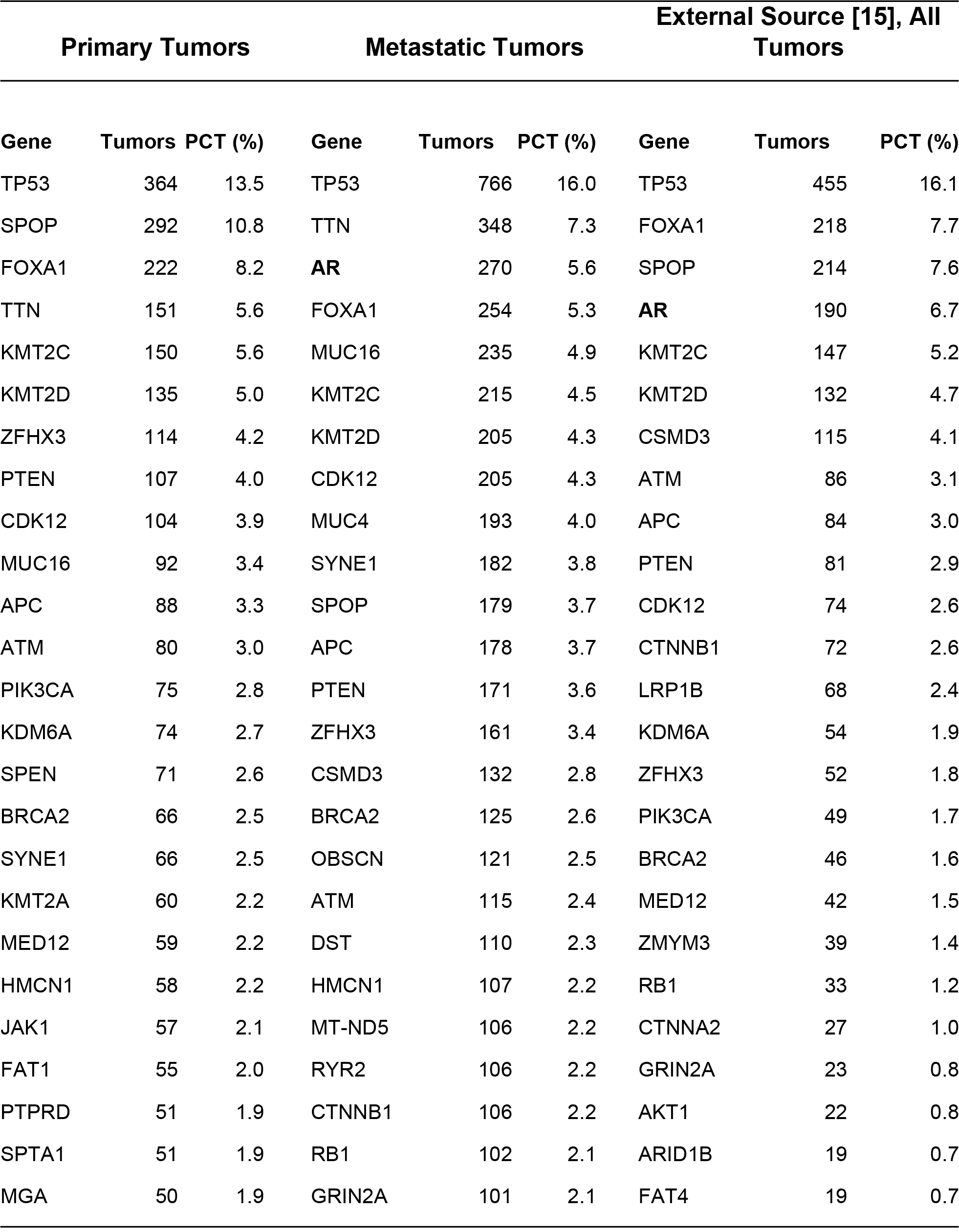
Frequency of mutations of the top 25 most mutated genes in PRAD.

| Primary Tumors |  |  | Metastatic Tumors |  |  | External Source [15], All Tumors |  |  |
| --- | --- | --- | --- | --- | --- | --- | --- | --- |
| Gene | Tumors | PCT (%) | Gene | Tumors | PCT (%) | Gene | Tumors | PCT (%) |
| TP53 | 364 | 13.5 | TP53 | 766 | 16.0 | TP53 | 455 | 16.1 |
| SPOP | 292 | 10.8 | TTN | 348 | 7.3 | FOXA1 | 218 | 7.7 |
| FOXA1 | 222 | 8.2 | **AR** | 270 | 5.6 | SPOP | 214 | 7.6 |
| TTN | 151 | 5.6 | FOXA1 | 254 | 5.3 | **AR** | 190 | 6.7 |
| KMT2C | 150 | 5.6 | MUC16 | 235 | 4.9 | KMT2C | 147 | 5.2 |
| KMT2D | 135 | 5.0 | KMT2C | 215 | 4.5 | KMT2D | 132 | 4.7 |
| ZFHX3 | 114 | 4.2 | KMT2D | 205 | 4.3 | CSMD3 | 115 | 4.1 |
| PTEN | 107 | 4.0 | CDK12 | 205 | 4.3 | ATM | 86 | 3.1 |
| CDK12 | 104 | 3.9 | MUC4 | 193 | 4.0 | APC | 84 | 3.0 |
| MUC16 | 92 | 3.4 | SYNE1 | 182 | 3.8 | PTEN | 81 | 2.9 |
| APC | 88 | 3.3 | SPOP | 179 | 3.7 | CDK12 | 74 | 2.6 |
| ATM | 80 | 3.0 | APC | 178 | 3.7 | CTNNB1 | 72 | 2.6 |
| PIK3CA | 75 | 2.8 | PTEN | 171 | 3.6 | LRP1B | 68 | 2.4 |
| KDM6A | 74 | 2.7 | ZFHX3 | 161 | 3.4 | KDM6A | 54 | 1.9 |
| SPEN | 71 | 2.6 | CSMD3 | 132 | 2.8 | ZFHX3 | 52 | 1.8 |
| BRCA2 | 66 | 2.5 | BRCA2 | 125 | 2.6 | PIK3CA | 49 | 1.7 |
| SYNE1 | 66 | 2.5 | OBSCN | 121 | 2.5 | BRCA2 | 46 | 1.6 |
| KMT2A | 60 | 2.2 | ATM | 115 | 2.4 | MED12 | 42 | 1.5 |
| MED12 | 59 | 2.2 | DST | 110 | 2.3 | ZMYM3 | 39 | 1.4 |
| HMCN1 | 58 | 2.2 | HMCN1 | 107 | 2.2 | RB1 | 33 | 1.2 |
| JAK1 | 57 | 2.1 | MT-ND5 | 106 | 2.2 | CTNNA2 | 27 | 1.0 |
| FAT1 | 55 | 2.0 | RYR2 | 106 | 2.2 | GRIN2A | 23 | 0.8 |
| PTPRD | 51 | 1.9 | CTNNB1 | 106 | 2.2 | AKT1 | 22 | 0.8 |
| SPTA1 | 51 | 1.9 | RB1 | 102 | 2.1 | ARID1B | 19 | 0.7 |
| MGA | 50 | 1.9 | GRIN2A | 101 | 2.1 | FAT4 | 19 | 0.7 |

The most commonly mutated genes are: TP53, SPOP, FOXA1, AR, PTEN, TTN, KMT2C, KMT2D, BRCA2, and others. On this list, TP53 is found to be the most highly mutated gene. It is known that TP53 has the most frequent alterations in all cancer types [16][17][18]. Being a common denominator, TP53 was excluded from this research.

The last set of mutations in Table 2 includes both primary and metastatic tumors. An important gene, Androgen Receptor (AR), appears highly mutated on this set and in our metastatic tumors list, but is not highly ranked in our primary tumors list. This is consistent with the fact that AR mutations are usually found to increase with cancer stage [19]. The androgen receptor (AR) has been used as the target for androgen deprivation therapy to treat de novo or recurrent metastatic disease [20]. Although this therapy works for a while, inhibiting tumor growth, most patients eventually develop resistance, and the disease becomes castration-resistant prostate cancer. AR is mutated at different rates at different metastatic sites, and thus, it was included in the calculations and algorithm.

#### Metastatic Site Data Analysis

The 25 most highly mutated genes by metastatic site were obtained, resulting in a total of 124 different genes. This indicates the wide variety of most-ranked mutated genes across the 8 metastatic sites analyzed in this research. Genes that are not mutated in at least 3 tumors were excluded, resulting in some metastatic sites having fewer than 25 most frequently mutated genes.

We define the “rank” as the order in which a gene appears most frequently mutated, #1 being the highest mutated and #25 the least mutated.

In Table 3, it can be seen that gene FOXA1 has the highest rank (#1) in metastases to the bone and lungs, but in metastases to the liver, it is ranked #9. FOXA1 does not show up as one of the most-ranked genes in other metastatic sites.

**Table 3.**
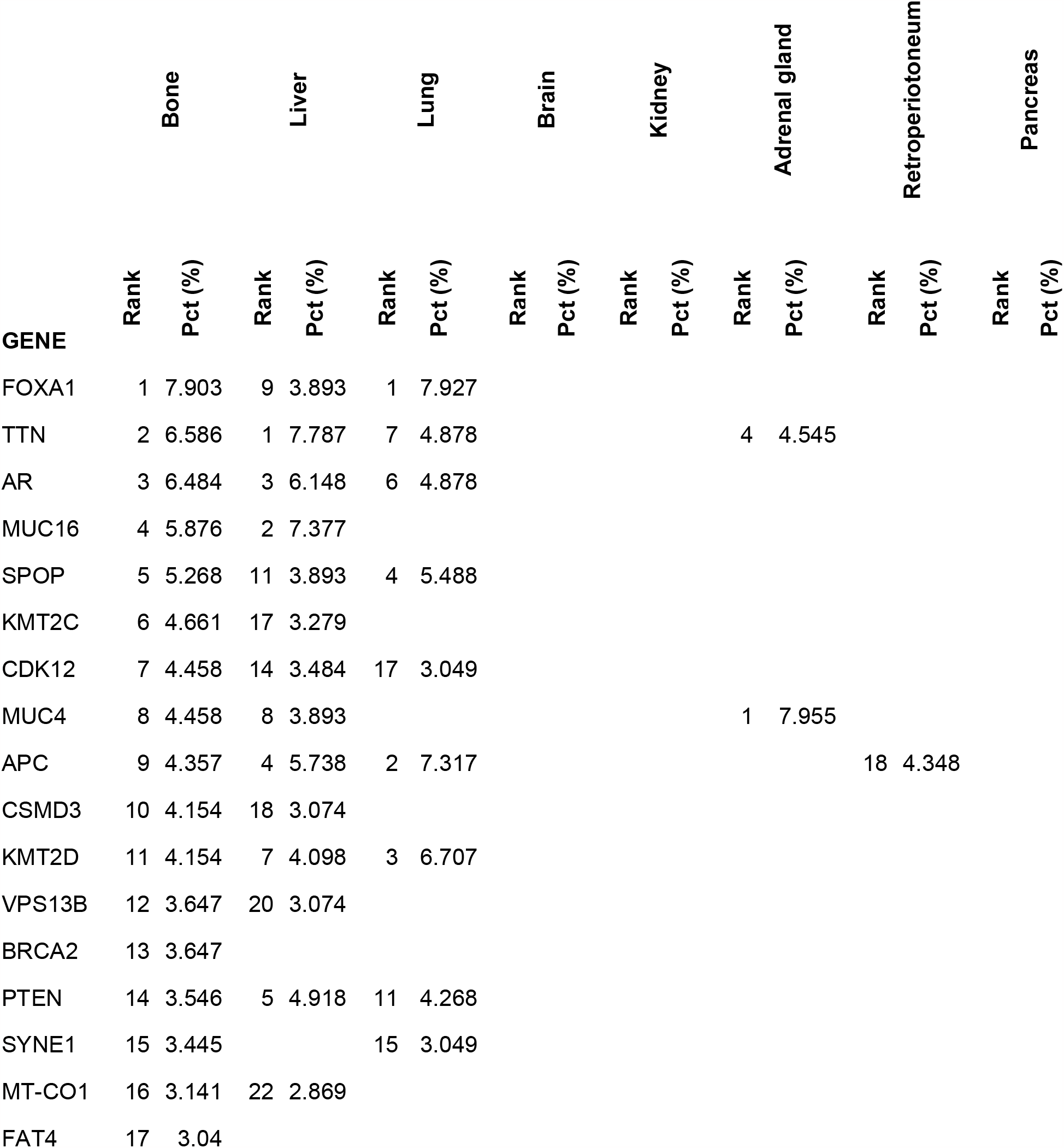
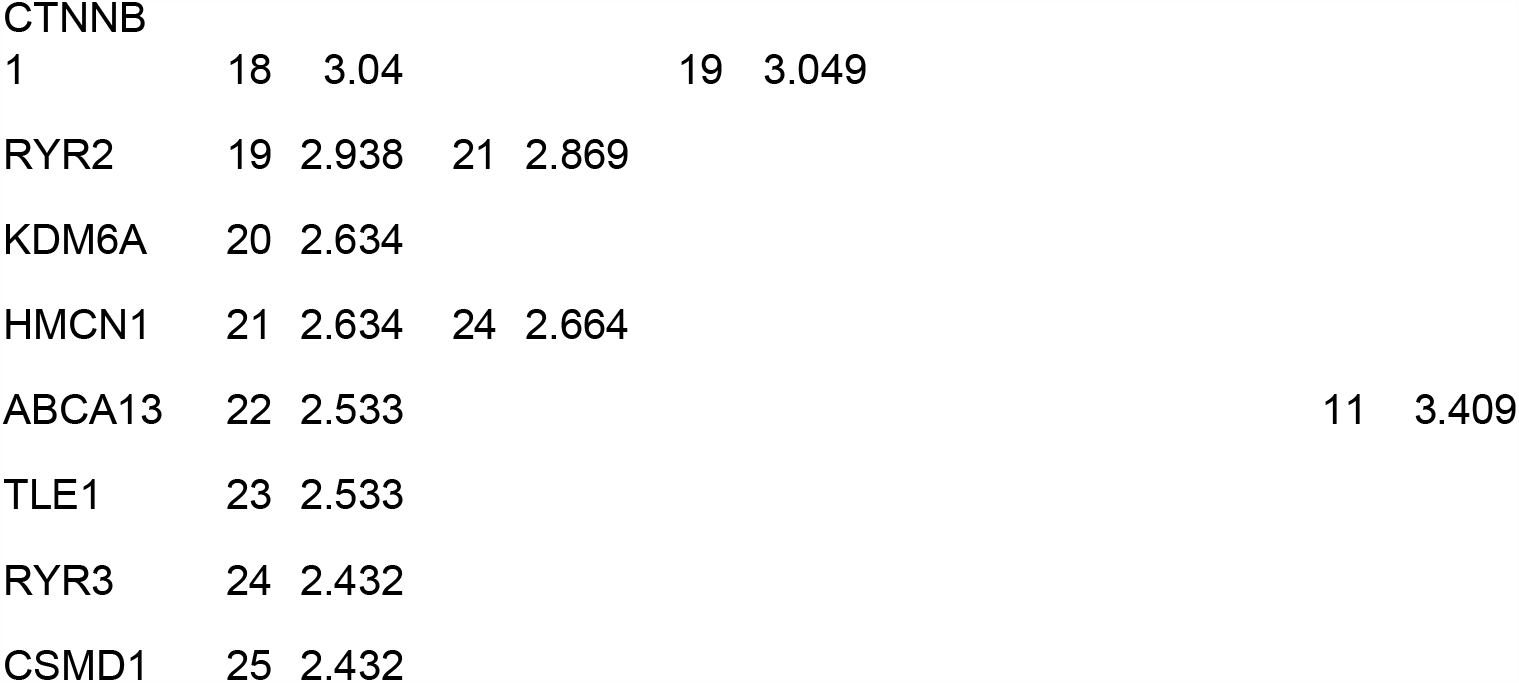
Mutational Landscape of Bone Metastasis vs. Other Metastatic Sites.

Bone and liver metastases have several common mutated genes, but at different rates of mutation. Bone and lung share less commonly mutated genes, and finally, bone metastases have no common mutated genes with the brain, kidney, and pancreas; only one common gene with retroperitoneum metastasis (APC); and only 3 common genes with adrenal gland metastases: TTN, MUC4, and ABCA13.

A similar analysis was done with all other metastatic sites, and it was found that the frequency of mutated genes among them is a feature that can be exploited as a pattern recognition variable. The lists of the top 25 most mutated genes in those 8 metastatic sites are greatly diverse.

Additionally, we found a list of 111 genes that are uniquely mutated at specific metastatic sites. These are genes that belong to the list of the top 25 most mutated genes. Metastases to the pancreas have the most number of unique mutated genes: 25. For this reason, even though we only have 5 cases of metastasis to the pancreas, it was possible to classify those tumors with good accuracy.

### The Prediction Algorithm

Based on our previous analysis, we designed an algorithm that, given a list of somatic mutations in a prostate cancer tumor, will predict the three most likely metastatic sites in order of probability.

The algorithm has 2 main steps:

1. Determine if the tumor has 4 or more genes that belong to the list of unique genes for a specific site. If that is the case, the metastatic site is predicted, and no further analysis is done.
2. If the metastatic site cannot be predicted by the uniqueness of its genes, a list of 8 scores, one per metastatic site, is calculated. For each gene in the tumor, the score of each metastatic site is calculated as the sum of the multiplication of each gene’s percentage within the MS times the percentage of the MS. This is a competition algorithm based on the variability of the weights of metastatic sites and genes within its MS.
3. At the end, the list is sorted by final score, and the three metastatic sites with the highest score are the answer.

## RESULTS AND DISCUSSION

The algorithm was implemented with code developed in T-SQL (Microsoft SQL Server 2022). After executing the algorithm on the data set of metastatic prostate tumors, it was found that we can predict metastatic sites with the following accuracies: bone (90.9%), retroperitoneum (87.5%), liver (83.0%), kidney (80.0%), pancreas (80.0%), adrenal gland (75.0%), brain (72.5%), and lung (71.1%), shown on Table 4.

**Table 4.** Prediction of Metastatic Site in Prostate Cancer Accuracy.

| Metastatic site | Cases (n) | Accuracy |
| --- | --- | --- |
| Bone | 506 | 90.9% |
| Liver | 218 | 83.0% |
| Lung | 76 | 71.1% |
| Brain | 16 | 72.5% |
| Kidney | 10 | 80.0% |
| Adrenal Gland | 8 | 75.5% |
| Retroperitoneum | 8 | 87.5% |
| Pancreas | 5 | 80.0% |

The accuracy was calculated after running the algorithm on all 847 metastatic tumors for which the metastatic site is known. The average accuracy was 80%.

The top three metastatic sites, bone, liver, and lung, are predicted mainly by the large amount of cases and, thus, their large weight in the competition formula to compute the score. The remaining sites, for which a few cases exist, are mainly predicted based on the number of unique genes that are on their uniqueness lists.

## CONCLUSIONS

We successfully developed a method and an algorithm that predict the most likely metastatic site of a primary prostate cancer tumor based on its somatic gene mutations. The accuracy of the predictions for eight metastatic sites ranges from 71.1% to 90.9%, with an average of 80.0%.

This research was aimed at developing a computational tool that could predict a metastatic growth even in the absence of clinical evidence at the time of RP and initial WB-MRI. All metastatic cancers start by shedding circulating tumor cells (CTC) into the circulation. The CTCs that survive the harsh environment of the circulation eventually extravasate to distant organs, giving rise to micrometastases, which in their initial stage cannot be imaged, and thus a false-negative report is presented.

Having a technological tool that warns of the probable presence of metastatic lesions, especially in organs where metastasis is not frequent, serves as an important guide in the treatment of patients with advanced PCa, who, based on the predicted metastases, would be monitored regularly and specifically.

## Data Availability

All data produced are available online at prostate.nano-bio-tek.com

https://prostate.nano-bio-tek.com

## DATA SHARING

A portal on our institution’s website was developed to share the results of this research. The prediction tool is freely available to all researchers, clinicians, and the general public. The URL to the portal is: https://prostate.nano-bio-tek.com/

Figure 3 shows an example of the results obtained by entering the mutated genes of a real tumor that metastasized to the lungs. The sample ID on CbioPortal.org is “P-0021894-T02-IM6” and additional information is freely available online on that website. For reference, the complete list of mutated genes is as follows:

**Figure 3.**
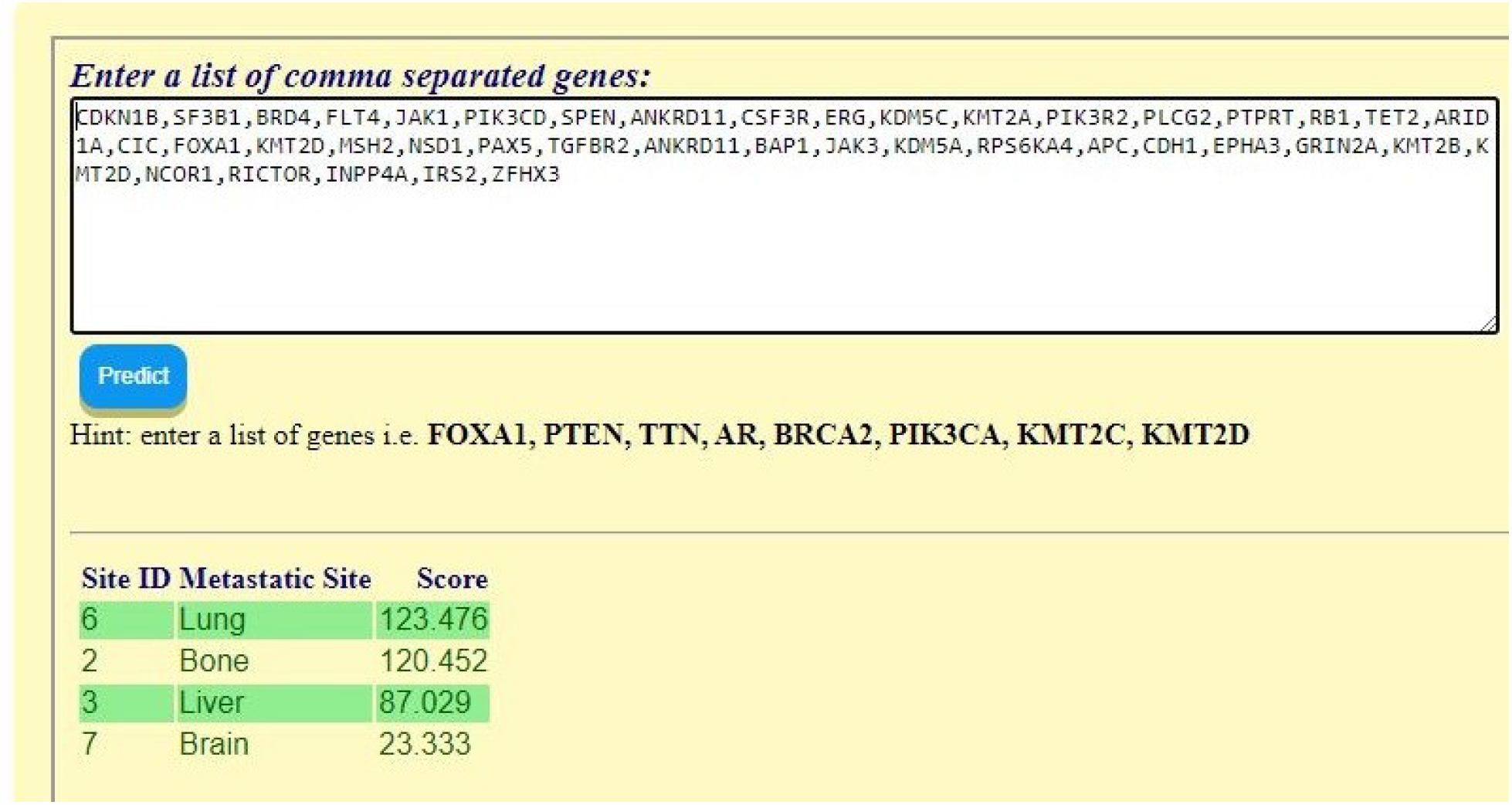
Prediction Tool Web Portal

CDKN1B,SF3B1,BRD4,FLT4,JAK1,PIK3CD,SPEN,ANKRD11,CSF3R,ERG,KDM5C,KMT2A,PI K3R2,PLCG2,PTPRT,RB1,TET2,ARID1A,CIC,FOXA1,KMT2D,MSH2,NSD1,PAX5,TGFBR2,ANKRD11,BAP1,JAK3,KDM5A,RPS6KA4,APC,CDH1,EPHA3,GRIN2A,KMT2B,KMT2D,NCOR1,RICTOR,INPP4A,IRS2,ZFHX3.

